# A Structure-Aware Self-Supervised Framework for Transferable Fetal Ultrasound Screening

**DOI:** 10.64898/2026.09.21.26361576

**Authors:** Yantong Zhu, Jiang Wu, Lan Gao, Na Zhang, Lan Wu, Hezhou Li, Kuifang Chen, Hong Luo, Congxin Sun, Daru Lu, Qingqing Wu, Hua Chen

## Abstract

Accurate prenatal screening from ultrasound imaging is severely constrained by the extreme spatial heterogeneity of fetal structures and the data scarcity inherent to rare congenital anomalies. To overcome these computational bottlenecks, we present RadarMAE, a data-efficient self-supervised representation learning framework tailored for fetal ultrasound analysis. By employing a novel, structure-aware masking strategy, RadarMAE enables robust pre-training on highly limited clinical datasets. Benchmarking the framework on first-trimester screening for trisomy 21 (Down syndrome), our model achieved exceptional diagnostic performance on an internal cohort, with an area under the curve (AUC) of 0.99, 97% accuracy, 94% sensitivity, and 98% specificity. Crucially, through domain-adversarial adaptation, the framework demonstrates robust cross-institutional generalization under extreme source-target data imbalance, bounding performance degradation to ***≤* 0.04** across all key metrics in external testing. Furthermore, by integrating spatial registration with explainable AI, we constructed the first population-level facial risk atlas for trisomy 21. This data-driven interpretability framework validates standard clinical markers while revealing the comparable diagnostic weight of underutilized features, such as mandibular dysmorphology. Ultimately, this approach provides a scalable, transparent, and high-performance solution for equitable prenatal diagnostics and phenotypic biomarker discovery.

## 1 Introduction

Prenatal screening is essential for identifying fetal congenital anomalies early in pregnancy, enabling timely clinical interventions that mitigate birth defects and safeguard maternal health. Currently, non-invasive screening relies on two complementary modalities: ultrasonography and cell-free DNA-based non-invasive prenatal testing (NIPT). While NIPT offers exceptional accuracy for common aneuploidies—achieving detection rates up to 100% and specificities near 99.8%—its clinical utility is constrained by a multi-day turnaround time, high cost, and dependence on centralized laboratory infrastructure. These barriers heavily restrict its widespread adoption, particularly in low- and middle-income countries.

Consequently, ultrasonography remains the cornerstone of global prenatal monitoring due to its real-time imaging capabilities, cost-effectiveness, and point-of-care safety. In mid-to-late gestation, structural anomalies associated with chromosomal disorders often become visually apparent; for instance, the absence of the nasal bone or an abnormal thickening of the nuchal translucency (NT) serve as classical ultrasound markers for Down syndrome (trisomy 21). However, waiting until the second trimester carries severe physical risks and psychological trauma associated with mid-trimester induction of labor. Shifting detection to the first trimester is a clinical imperative, yet early-stage ultrasound screening remains highly ineffective. First-trimester manual screening yields a detection rate of only 69% (with *∼*92% specificity), compared to approximately 90% (*∼* 95% specificity) in the second trimester. This disparity stems from the fact that early-stage ultrasound images suffer from low resolution, acoustic artifacts, and highly variable structural features, making quantitative measurements exceptionally vulnerable to operator subjectivity and variations in scanning angles.

Deep learning, particularly self-supervised learning (SSL), offers a promising pathway to overcome these manual limitations by automatically extracting robust representations from complex medical images without requiring exhaustive expert labeling. While SSL has driven breakthroughs in oncology and ophthalmology, adapting these methods to fetal ultrasound analysis presents severe computational bottlenecks. Standard SSL frameworks often rely on large-scale pre-training datasets containing millions of natural or medical images to build generalized feature hierarchies. Fetal ultrasound datasets, by contrast, are inherently sparse, localized, and highly heterogeneous. Significant domain shifts exist between different imaging devices, and data quality varies drastically across clinical institutions. Furthermore, the “black-box” nature of traditional deep networks severely impedes clinical trust. In high-stakes prenatal diagnostics, where algorithmic errors directly alter pregnancy management, there is an urgent need for sample-efficient, domain-robust, and highly interpretable AI systems. To address these fundamental limitations, here we introduce a self-supervised fetal ultrasound representation learning framework specifically engineered for sample-efficient, heterogeneous prenatal imaging, benchmarked on first-trimester screening for trisomy 21 (Down syndrome). Our framework overcomes the data-scarcity bottleneck of traditional self-supervised learning through a novel radar-shaped masked autoencoder, which yields robust feature representations from limited clinical datasets by optimizing the perception of complex fetal facial structures. To mitigate severe domain shifts across clinical environments, we implement a cross-institutional adaptation scheme that preserves diagnostic efficacy when transferred to resource-limited primary healthcare facilities, even in extreme cases of local data scarcity. Finally, to establish clinical trust, we move beyond single-instance post-hoc explanations by integrating fetal image registration directly with model interpretation. By mapping and aggregating maximum network activation coordinates into a standardized facial canonical space, our framework constructs a population-wide, disease-level facial risk atlas for trisomy 21. Together, these algorithmic advancements demonstrate excellent cross-device generalization and provide an interpretable, scalable path toward equitable, real-time clinical deployment in prenatal care. The overall study workflow is shown in Figure 1.

**Fig 1.**
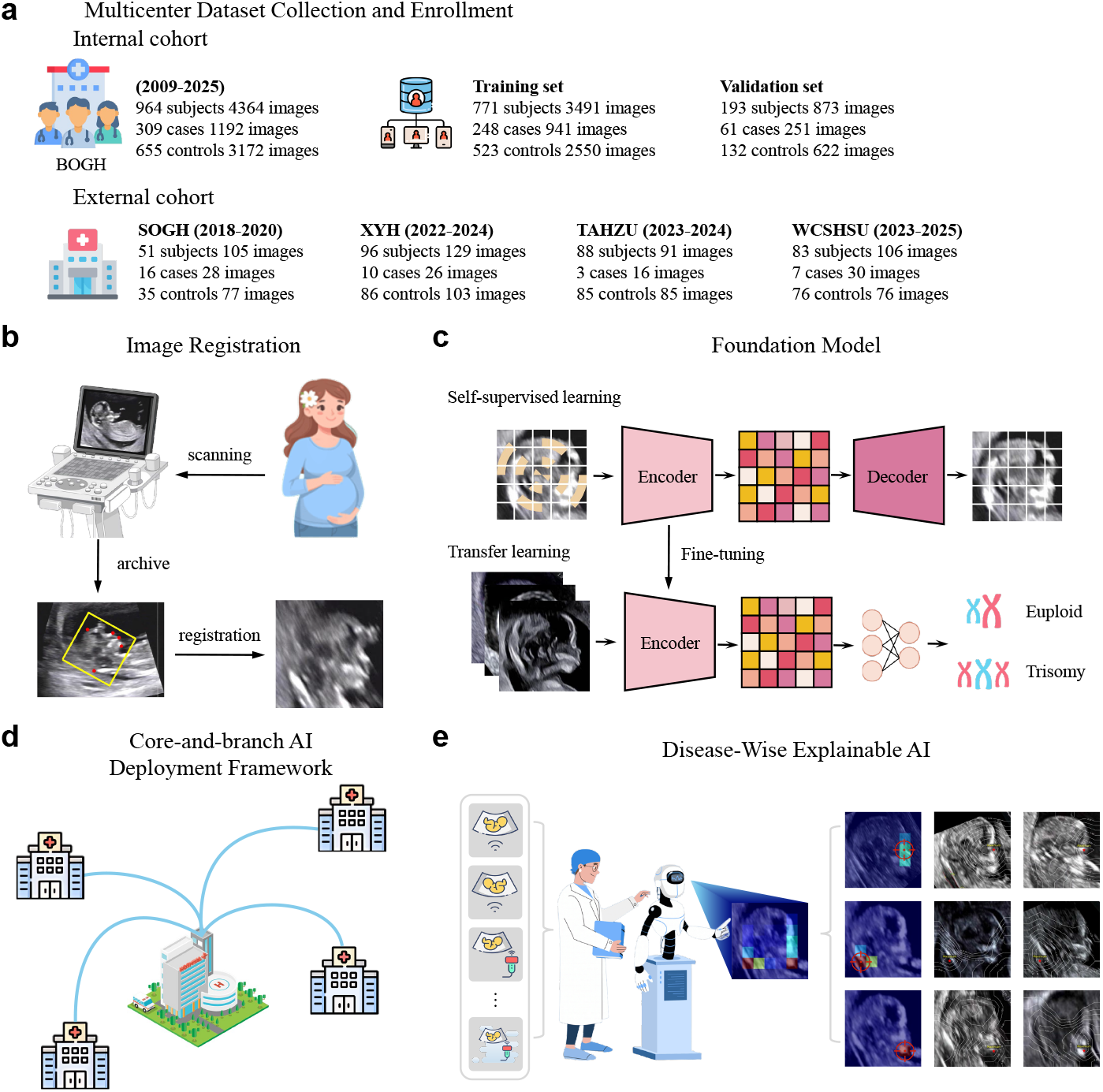
Study workflow. (a) Multicenter Dataset Collection and Enrollment: BOGH images are stratified into training (80%) and validation (20%) cohorts; XYH, SOGH, TAHZU, and WCSHSU images are used as cross-institutional external validation. (b) Standardized Spatial Registration: Retrospectively collect ultrasound images of the midsagittal plane, undergo image registration, and ROI cropping. (c) Self-Supervised Representation Learning: Pre-training through a radar-shaped mask, followed by fine-tuning for trisomy 21 screening. (d) Illustration of the core-and-branch AI deployment framework. (e) Disease-wise explainable AI for the risk atlas generation.

## 2 Results

### 2.1 System Overview and Standardized Spatial Registration

To enable automated, high-precision analysis of fetal ultrasound images, we first established an end-to-end processing pipeline that standardizes the spatial representation of craniofacial structures. While established clinical markers for trisomy 21—such as nuchal translucency (NT) and nasal bone hypoplasia—are highly localized [1–3], raw ultrasound data exhibit extreme spatial variability. Conventional region-of-interest (ROI) cropping typically relies on simple bounding boxes around the fetal head [4, 5]. However, this approach addresses only basic coordinate translation; it fails to correct for severe rotational variances (head pose) and laterality (left-right flipping symmetry), which structurally misalign the data and confound downstream neural network feature extraction.

To overcome this, we developed a standardized image registration process that maps all images into a structurally consistent coordinate space [6]. As illustrated in Figure 2a, our scheme identifies five persistent craniofacial reference points: the external occipital region (EO), head crown (HC), nasal tip (NAT), alveolar ridge (AR), and jaw (JAW) [7]. These specific landmarks were selected for their positional stability throughout gestation and high annotation reproducibility across sonographers, thereby minimizing inter-operator variability. Using these landmark pairs, we estimate a transformation matrix to normalize translation, rotation, and horizontal flipping across the entire dataset.

**Fig 2.**
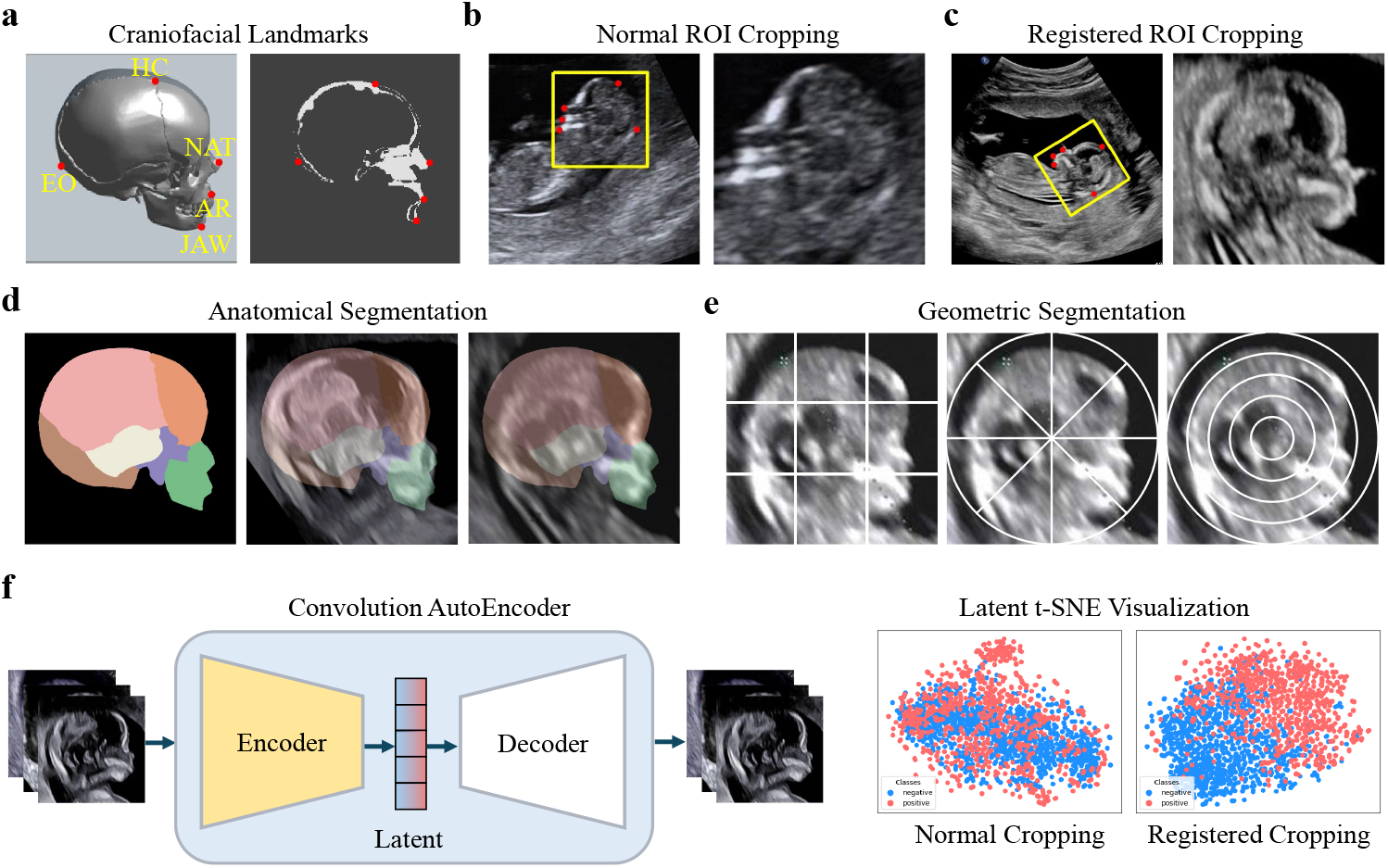
System Overview and Standardized Spatial Registration (a) Structural definition of five craniofacial reference points: the external occipital region (EO), head crown (HC), nasal tip (NAT), alveolar ridge (AR), and jaw (JAW). (b) Normal ROI cropping limit to translation transformation. Proposed registered ROI cropping, additional spatial transformation: rotation, horizontal flipping. and (e) The registered coordinate system ensures spatial consistency for flexible segmentation approaches - structural guidance or configurable geometric primitives - facilitating precise facial substructure extraction. (f) Convolution autoencoder and t-SNE are employed for latent space visualization, where red and blue scatter points represent trisomy 21 and euploid control cases, respectively. The registered cropping yields a clearer case-control separation compared to unprocessed data.

This registered ROI cropping provides a critical structural advantage over conventional methods (Figure 2b,c). By enforcing strict spatial correspondence, our normalization ensures universal template applicability [8], allowing standardized segmentation masks or flexible superpixel primitives to be applied consistently across all images without spatial misalignment [9] (Figure 2d,e).

Most importantly, this spatial registration effectively isolates the true diagnostic signals from ubiquitous ultrasound background noise, hardware limitations, and dynamic occlusions (e.g., umbilical cord or limbs). To quantitatively validate this, we evaluated the latent space representations of both raw and registered images using an unsupervised convolutional autoencoder mapped via t-SNE [10]. As shown in Figure 2f, registered images produce significantly tighter intra-class clustering and enhanced inter-class separation between euploid controls and trisomy 21 cases. This empirically confirms that normalizing spatial variance successfully projects the raw ultrasound data into a highly discriminative feature space, providing a robust, noise-reduced basis for downstream self-supervised representation learning.

### 2.2 Self-Supervised Representation Learning

The development of diagnostic AI models for prenatal screening is severely constrained by data scarcity. Due to the low incidence of rare genetic diseases (trisomy 21 incidence is < 0.1%), primary healthcare facilities rarely encounter enough positive cases—often fewer than 10 annually—to support robust supervised learning. These limitations restrict the validation performance of research prototypes and compromise cross-institutional deployment, impeding the clinical translation of medical AI.

While self-supervised learning (SSL) mitigates data scarcity in other domains by deriving supervisory signals directly from unannotated data, standard SSL frameworks struggle when applied to fetal ultrasound. By systematically masking random image patches and reconstructing the missing content, masked representation learning has driven breakthroughs in natural images [11], histopathology [12], and retinal imaging [13]. However, this efficacy relies heavily on abundant spatial redundancy. Fetal ultrasound images, by contrast, exhibit extreme spatial heterogeneity. Diagnostically critical features, such as nuchal translucency and specific craniofacial structures, are densely packed into minute structural regions, whereas surrounding areas contain sparse information. Standard random-masking strategies risk completely occluding these crucial regions, preventing the model from learning clinically meaningful representations.

To overcome this structural bottleneck, we propose an efficient self-supervised framework tailored specifically to fetal facial topologies. Rather than employing random patches, our method systematically masks image sectors at controlled radial and angular intervals. As illustrated in Figure 3 (a) and (b), concentric circular masking forces the network to learn the continuous curvature of craniofacial structures (e.g., cranial bones), while angular sector masking enhances the perception of elongated morphologies (e.g., the mandible). By synergistically integrating these patterns—mimicking a radar sweep (Figure 3 (c))—this structured approach preserves sufficient neighboring context for accurate reconstruction. Consequently, we designate this architecture the Radar Masked Autoencoder (RadarMAE), which actively compels the network to learn precise, localized structural representations even from strictly limited datasets.

**Fig 3.**
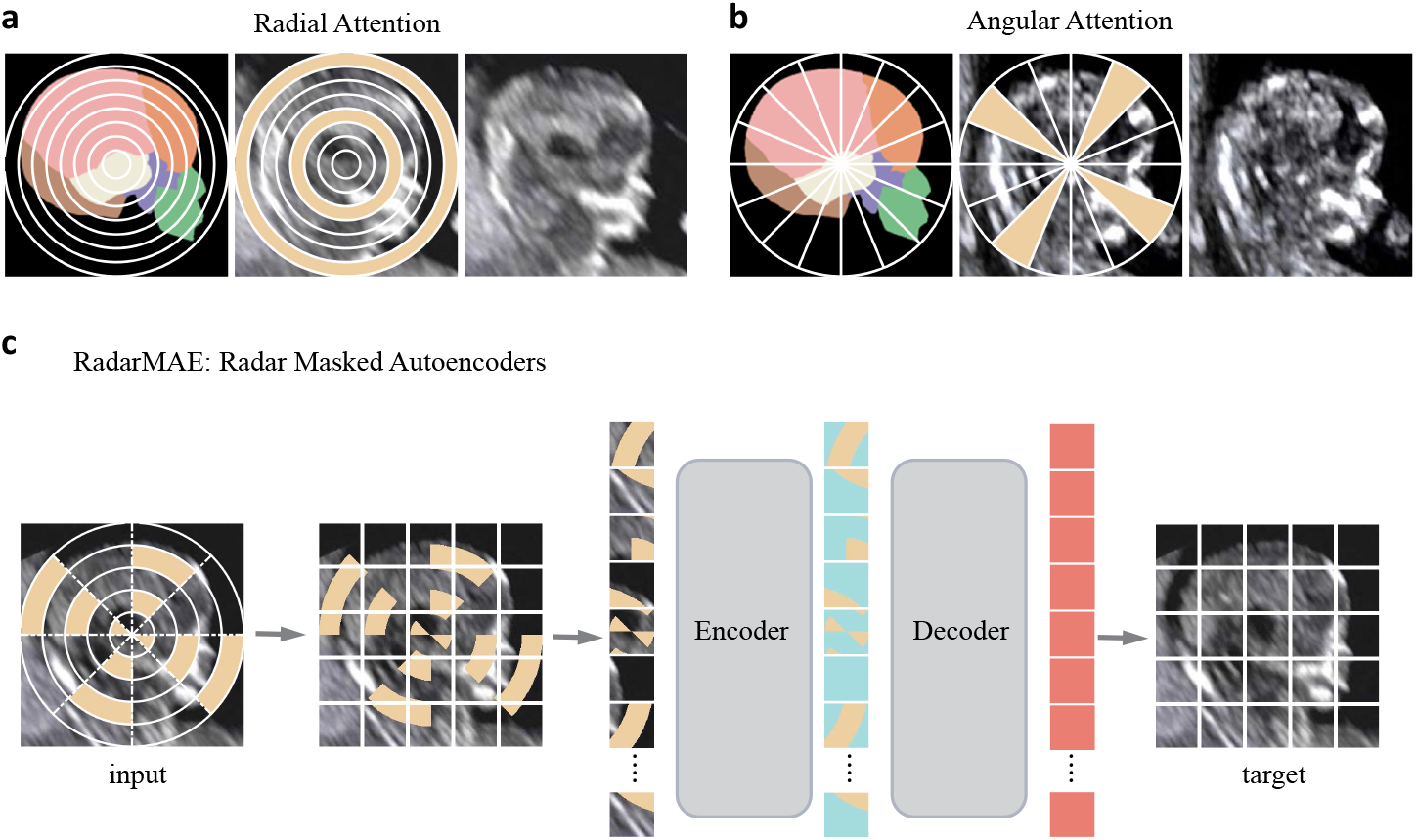
RadarMAE Self-Supervised Architecture (a) Radial Attention: Masking concentric rings at regular intervals along the radius forces focus on curved craniofacial structures. (b) Angular Attention: Masking sectors at angular intervals prevents complete occlusion of elongated morphology. Each triplet shows: the division of the masked pattern (left), the masked image (middle), and the corresponding ground-truth image (right). (c) In our RadarMAE architecture, we perform pixel-level occlusion on input images. Each encoder block can receive a variable number of pixel-level masks. The encoder and decoder maintain identical token counts. Following pre-training, we discard the decoder and utilize the encoder on uncorrupted images for downstream tasks.

To evaluate the efficacy of RadarMAE, we first established a strong baseline using conventional Convolutional Neural Networks (CNNs). When combined with our spatial registration, a transfer-learned ResNet achieved an AUC of 0.98, an accuracy of 0.95, a sensitivity of 0.90, and a specificity of 0.96, confirming the synergistic benefits of spatial normalization and pre-training for CNNs. Vision Transformers (ViTs) conventionally underperform CNNs in small-data regimes [14] due to a lack of inductive biases. However, pre-training the ViT architecture with our customized RadarMAE task fundamentally reversed this limitation. By explicitly enhancing the perception of fetal facial structures during the self-supervised reconstruction phase, the ViT + RadarMAE framework achieved superior diagnostic performance across all metrics (AUC: 0.99, accuracy: 0.97, sensitivity: 0.93, specificity: 0.98), effectively surpassing the optimized CNN baseline. Figure 4 and Table 1 summarize the performance metrics across all model configurations.

**Table 1.** Performance of different model settings. All models are trained on the internal cohort and evaluated on the internal testing cohort.

| Model Performance | Measure (95% CI) |  | Sensitivity | Specificity |
| --- | --- | --- | --- | --- |
|  | AUC | Accuracy |  |  |
| Convolutional Neural Network |  |  |  |  |
| ResNet + Normal + Scratch | 0.90 (0.88-0.92) | 0.90 (0.88-0.92) | 0.82 (0.77-0.86) | 0.94 (0.92-0.96) |
| ResNet + Normal + Pretrained | 0.98 (0.97-0.99) | 0.94 (0.93-0.96) | 0.89 (0.85-0.92) | 0.96 (0.95-0.98) |
| ResNet + Registered + Scratch | 0.97 (0.96-0.98) | 0.92 (0.90-0.94) | 0.88 (0.85-0.92) | 0.93 (0.91-0.95) |
| ResNet + Registered + Pretrained | 0.98 (0.97-0.99) | 0.95 (0.93-0.96) | 0.90 (0.86-0.93) | 0.96 (0.95-0.98) |
| Vision Transformer |  |  |  |  |
| ViT + Scratch | 0.97 (0.96-0.98) | 0.93 (0.92-0.95) | 0.82 (0.77-0.87) | 0.98 (0.97-0.99) |
| ViT + Pretrained | 0.97 (0.96-0.98) | 0.94 (0.92-0.95) | 0.86 (0.82-0.90) | 0.97 (0.95-0.98) |
| ViT + MAE | 0.98 (0.98-0.99) | 0.94 (0.93-0.96) | 0.86 (0.82-0.90) | 0.98 (0.97-0.99) |
| ViT + RadarMAE | 0.99 (0.98-0.99) | 0.97 (0.96-0.98) | 0.93 (0.90-0.96) | 0.98 (0.97-0.99) |

**Fig 4.**
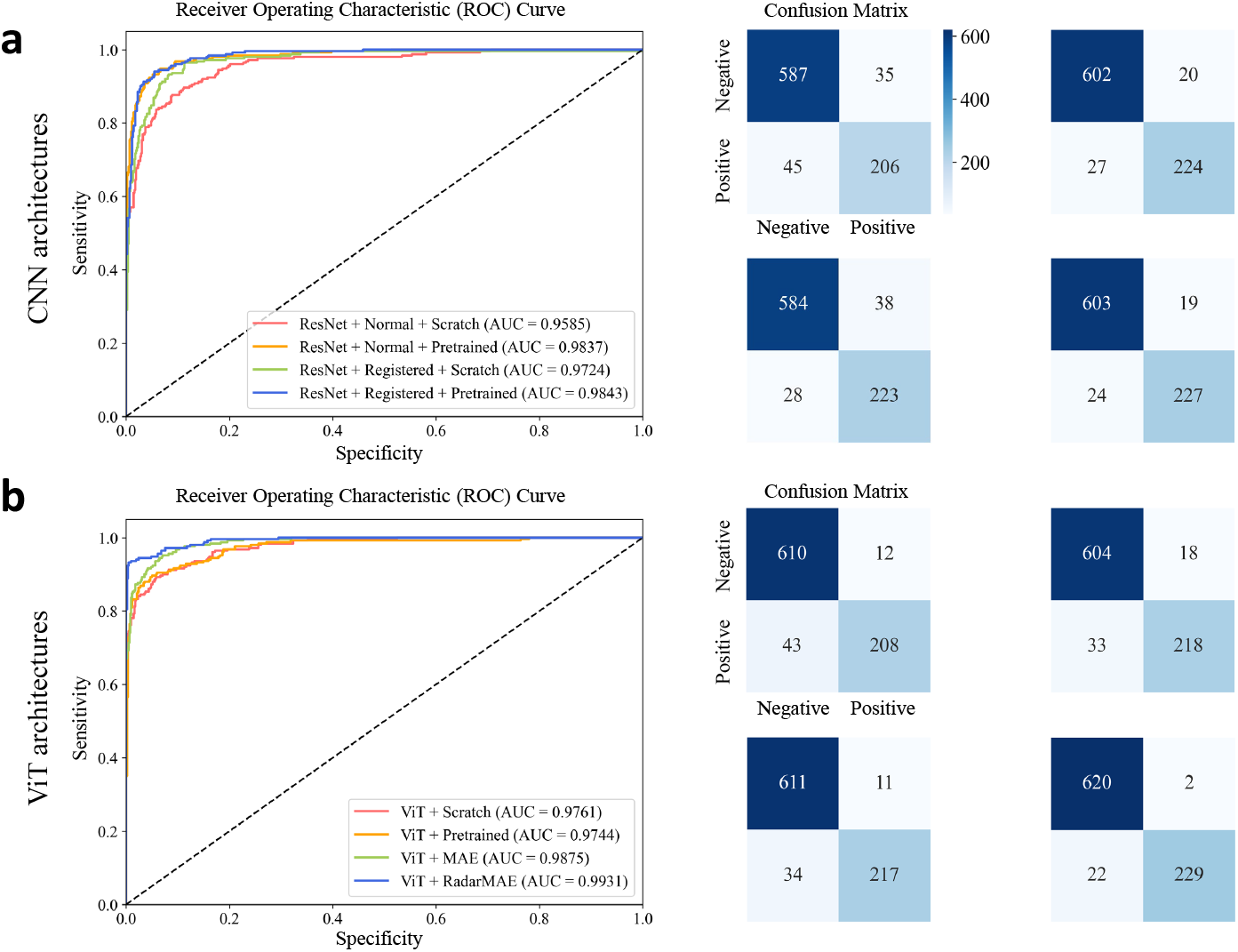
Performance Evaluation of ROC Curves and Confusion Matrices. (a) and (b) comparative analysis of CNN versus ViT architectures. For each architecture, the left panel displays ROC curves while the right panel presents corresponding confusion matrices. The quadrant-arranged confusion matrices (upper left, upper right, lower left, lower right) correspond to the red, orange, green, and blue model configurations indicated in the ROC curves.

### 2.3 Core-and-Branch AI Deployment Framework

Cross-institutional deployment of medical AI models frequently fails due to domain shifts, extreme target-domain data scarcity, and a lack of local annotations. To overcome these translational barriers, we developed a “Core-and-Branch” domain-adversarial framework that enables robust deployment under zero-label-leakage conditions. This dual-branch architecture pairs a primary diagnostic classifier with an adversarial domain classifier, forcing the network to project both source and target images into a unified, institution-agnostic feature space. We deployed the selfsupervised RadarMAE encoder as the feature extractor, utilizing the labeled BOGH cohort as the source domain for training and internal validation, while four independent clinical cohorts (XYH, SOGH, TAHZU, and WCSHSU) served as external target domains. During domain adaptation, the model accessed target-domain images exclusively without diagnostic labels, providing a stringent, real-world assessment of cross-institutional generalization.

The framework demonstrated exceptional robustness across diverse clinical settings. In the BOGH internal validation cohort, the model achieved an AUC of 0.99, an accuracy of 0.97, a sensitivity of 0.94, and a specificity of 0.98, successfully identifying 58 of 61 (95%) confirmed trisomy 21 cases. Crucially, this diagnostic efficacy was preserved across the four external testing cohorts, maintaining an aggregate AUC of 0.98, an accuracy of 0.95, a sensitivity of 0.93, and a specificity of 0.94. Notably, the framework detected 100% of trisomy 21 cases in both the XYH and TAHZU external cohorts. These results establish that our AI-augmented ultrasound pipeline can approach the diagnostic sensitivity of cell-free DNA (cfDNA) screening, while retaining the critical advantages of real-time, point-of-care availability and economic viability in resource-limited environments (Table 2, Figure S1).

**Table 2.** Comparisons of Model Screening Performance with state-of-the-art methods. Cross-domain (CD) means that the training and testing sets come entirely from different domains. ‘-’ indicates data that is not reported.

| Model Performance | Measure (95% CI) |  | Accuracy | Sensitivity | Specificity |
| --- | --- | --- | --- | --- | --- |
|  | CD | AUC |  |  |  |
| SOTA Method |  |  |  |  |  |
| Sun et al. [15] | ✗ | 0.97 (0.96-0.99) | 0.92 (0.86-0.97) | - | - |
| Zhang et al. [4] | ✗ | 0.95 (0.93-0.98) | 0.89 (0.84-0.92) | 0.78 (0.69-0.86) | 0.94 (0.89-0.97) |
| Tang et al. [5] | ✗ | - | 0.83 | 0.83 (0.51-0.97) | 0.94 (0.89-0.97) |
| Our Method |  |  |  |  |  |
| BOGH Internal Cohort | ✗ | 0.99 (0.98-0.99) | 0.97 (0.96-0.98) | 0.94 (0.92-0.96) | 0.98 (0.97-0.99) |
| XYH External Cohort | ✓ | 0.99 (0.99-1.00) | 0.97 (0.94-0.99) | 1.00 (1.00-1.00) | 0.96 (0.92-0.99) |
| SOGH External Cohort | ✓ | 0.98 (0.94-1.00) | 0.96 (0.92-0.99) | 0.93 (0.81-1.00) | 0.97 (0.93-1.00) |
| TAHZU External Cohort | ✓ | 0.98 (0.96-1.00) | 0.95 (0.90-0.99) | 1.00 (1.00-1.00) | 0.94 (0.88-0.99) |
| WCSHSU External Cohort | ✓ | 0.99 (0.98-1.00) | 0.98 (0.95-1.00) | 0.97 (0.89-1.00) | 0.99 (0.96-1.00) |

The success of this domain-adversarial adaptation is heavily dependent on the stability of the underlying feature representations. While adversarial training effectively aligns feature distributions across institutions, target-domain performance ultimately relies on the generalizability of the base extractor. Conventional architectures, including ResNet and standard MAE, exhibited severe instability in this deployment paradigm; they failed to achieve consistent convergence across domains, and in some configurations, the adversarial branch actively impaired the primary classifier, triggering training divergence. By contrast, because RadarMAE learns highly structured, structure-aware facial representations during its self-supervised pre-training, it provided a uniquely stable feature space for downstream alignment. As shown in Figure S1, RadarMAE achieved smooth, synchronized convergence in both loss and accuracy, bounding the performance degradation between internal and external cohorts to less than 0.03 across AUC, accuracy, and specificity metrics.

### 2.4 Disease-Wise Explainable AI

While nuchal translucency (NT) thickening has been the cornerstone sonographic marker for trisomy 21 screening, complementary first-trimester markers—such as nasal bone hypoplasia [16], widened inner canthal distance [17], and mandibular hypoplasia [18]—can significantly enhance detection rates and reduce false positives [19]. However, manual assessment of these craniofacial landmarks requires highly specialized sonographic expertise. Moreover, traditional clinical workflows rely on isolated, single-feature measurements, leaving the complex spatial relationships and combinatorial importance of these morphological markers largely underexplored. To characterize the spatial distribution and relative importance of disease-associated features in model predictions, we employed a data-driven interpretability framework that aggregates neural network activation patterns across images within a standardized population-level facial space.

As a baseline, Grad-CAM activation patterns for euploid controls exhibit diffuse, centrally distributed attention. The maximum heatmap intensity consistently localizes near the image centroid and decays uniformly toward the periphery (Figure 6 (b)). This diffuse activation confirms that the network identifies no overriding pathological visual preference in healthy fetuses.

**Fig 5.**
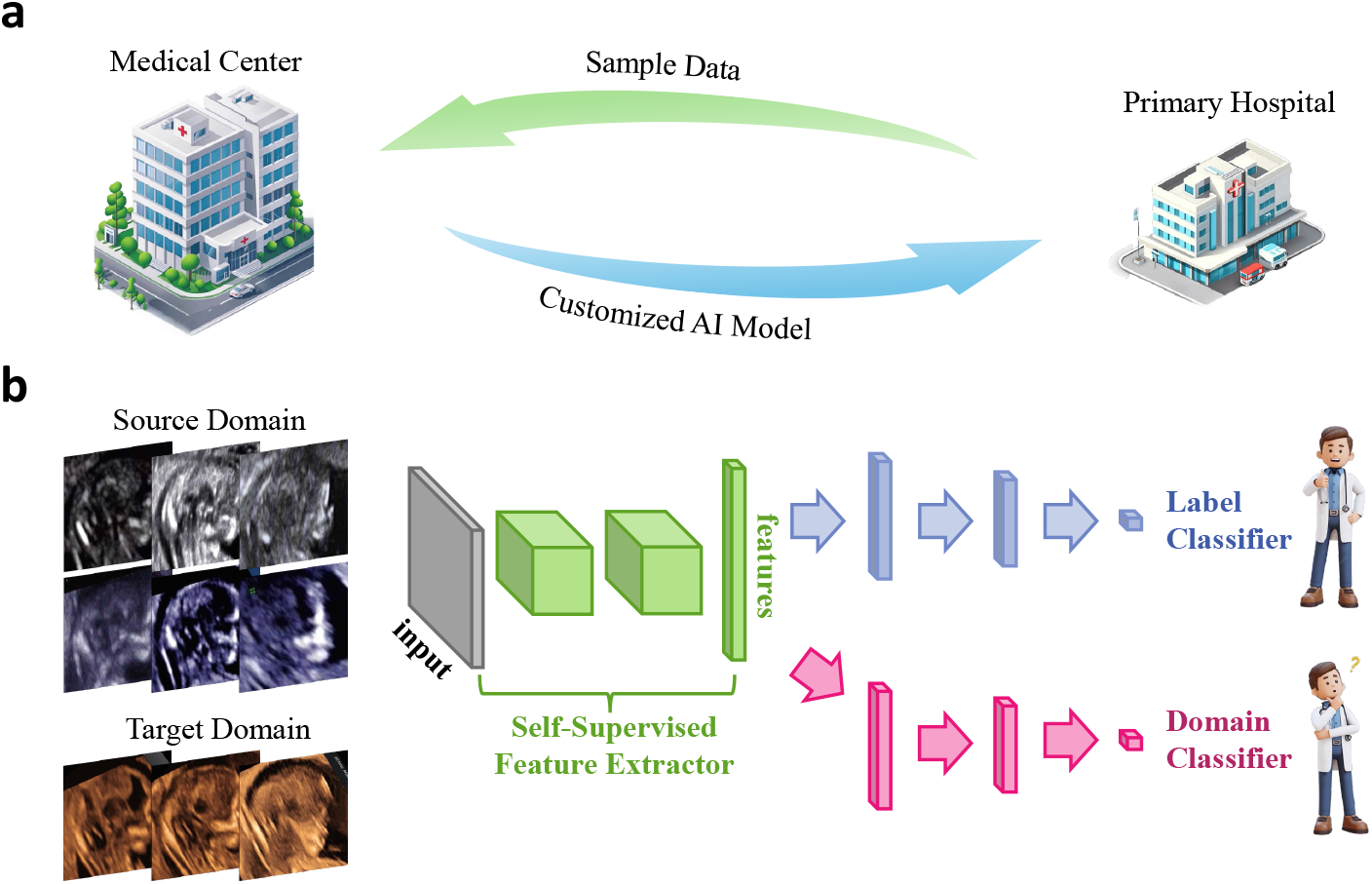
Core-and-branch AI Deployment Framework. (a) Centralized-Distributed Collaborative Paradigm: Coordination between top-level medical centers and primary care hospitals. (b) Our Model Architecture: Integration of RadarMAE feature extraction and domain-adversarial training optimization.

**Fig 6.**
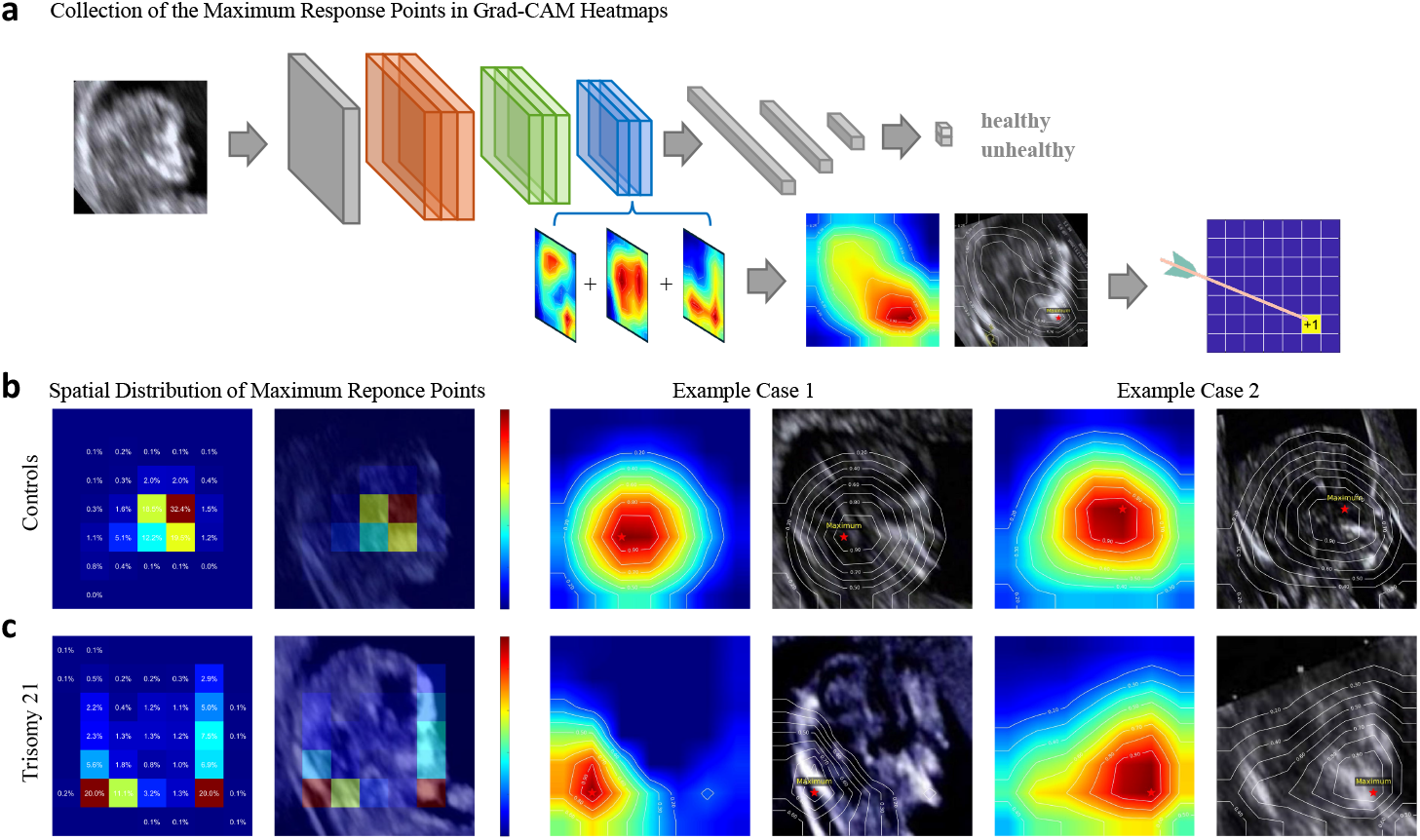
Population-Level Facial Risk Mapping: (a) Grad-CAM paired with the collection of maximum response points. (b) and (c) The spatial distribution of maximum response points for euploid controls and trisomy 21, accompanied by two representative cases for illustrative purposes.

In stark contrast, the maximum response distribution for trisomy 21 cases reveals a distinctly pathological morphological signature (Figure 6 (c)). The network aggressively targets the nuchal and mandibular regions, with each area independently accounting for approximately 20% of the aggregate visual focus. Crucially, while large-scale cohort studies and clinical guidelines [19] overwhelmingly rely on NT as the primary imaging criterion, mandibular abnormalities have received substantially less clinical priority. Our data-driven risk atlas demonstrates that the network weights mandibular hypoplasia as heavily as NT thickening. This suggests that current NT-centric protocols risk missing atypical cases, and that systematically integrating mandibular assessments into broad screening workflows could significantly enhance overall detection sensitivity.

Furthermore, multiple facial regions collectively accounted for approximately 20% of the model’s attention, including small craniofacial structures such as the eye sockets and nasal bones. In standard clinical practice, the small scale of these early-gestation structures presents severe technical barriers to reliable manual measurement. The AI’s ability to consistently capture the spatial and probabilistic correlations of these anterior abnormalities highlights their latent diagnostic value. By moving beyond isolated manual heuristics to a comprehensive morphological risk atlas, this framework not only independently validates established clinical markers but also exposes combinatorial phenotypic patterns that reflect the complex developmental disruptions underlying trisomy 21.

## 3. Discussion

This study presents a standardized, AI-driven ultrasound analysis framework designed to fundamentally enhance first-trimester prenatal screening. By eliminating the reliance on manual annotation and subjective heuristics, our approach overcomes critical clinical bottlenecks: profound dependencies on operator expertise, stringent time constraints during image acquisition, and the inherent difficulty of visually quantifying subtle craniofacial dysmorphology. Our multi-center evaluation confirms that the model’s robust screening performance generalizes effectively to resource-constrained healthcare settings. By lowering the expertise threshold required for early trisomy 21 detection, this framework facilitates scalable deployment across diverse clinical environments, directly promoting broader, more equitable access to high-quality prenatal care.

Beyond its immediate diagnostic utility, our framework advances explainable AI to construct the first population-level, disease-wise ultrasound risk atlas for trisomy 21. This atlas not only independently corroborates established landmark-based sonographic markers but also maps high-risk craniofacial regions—such as mandibular dysmorphology—that remain difficult to assess using conventional, single-feature clinical criteria. As an interpretable assistive tool, this mapping enhances expert judgment during first-trimester screening and supports data-driven biomarker discovery. Ultimately, this approach transitions fetal facial analysis from subjective, qualitative observation to objective, spatially resolved quantification, paving the way for more comprehensive non-invasive screening protocols.

Technically, our framework addresses the multifaceted challenges of medical AI deployment through three core innovations: a robust self-supervised RadarMAE encoder that enables sample-efficient representation learning, a domain-adversarial architecture that facilitates cross-institutional generalization without local annotations, and a disease-level interpretability mechanism that generates clinically actionable insights. Although validated here for trisomy 21, the proposed framework may be extended to other genetic disorders that manifest as detectable fetal morphological alterations. Nevertheless, we acknowledge several limitations. First, while our study encompasses diverse imaging hardware, the training and evaluation datasets are derived exclusively from tertiary care centers in China; establishing global applicability will require extensive external validation across diverse genetic populations and lower-tier clinical settings. Second, this study focuses strictly on trisomy 21, leaving the model’s efficacy for detecting other aneuploidies or isolated structural anomalies unexplored. Finally, although our explainable AI techniques successfully elucidate the model’s decision-making process, translating these population-level visual heatmaps into seamlessly integrated, real-time user interfaces for routine clinical workflows will require further ergonomic and software refinement.

## 4 Conclusion

We have developed a comprehensive, computationally efficient diagnostic framework that establishes a new paradigm for automated fetal ultrasound analysis. By utilizing a novel radar-shaped masked autoencoder, our method extracts highly robust, operator-independent phenotypic representations from inherently limited and spatially heterogeneous clinical datasets. Integrated with domain-adversarial adaptation, the model achieves state-of-the-art screening performance for trisomy 21 and demonstrates exceptional cross-institutional resilience, maintaining high accuracy even under extreme sample imbalances. Furthermore, by projecting network attention into a standardized facial canonical space, we expose complex pathogenic patterns of craniofacial anomalies that transcend traditional manual measurements. By bridging the gap between deep learning prototypes and practical clinical deployment, this technology provides an accessible, transparent, and high-performance screening solution poised to democratize early prenatal risk assessment on a global scale.

## 5 Methods

### 5.1 Multi-center Fetal Cohort Ultrasound Image Dataset

This study utilizes data collected by the ambispective cohort of China Common Early-Trimester Abnormalities. We collect all archived fetal ultrasound images from five tertiary hospitals in China. Two-dimensional midsagittal images of the fetal face are selected from singleton pregnancies at 11 to 13+6 weeks of gestation. The inclusion criteria are: (a) a fetal karyotype confirmed by chorionic villus sampling or amniocentesis; and (b) clear visualization of the nasal bone, frontal bone, overlying nasal skin, and the anterior margins of the maxilla and mandible. The dataset comprises 345 cases of trisomy 21 confirmed via invasive testing. We then match these with 937 euploid fetuses, randomly selected using the same imaging criteria applied to the trisomy 21 group.

All ultrasonographic data are acquired following the ISUOG nuchal translucency measurement guidelines. We obtain data from five tertiary prenatal diagnostic centers: Beijing Obstetrics & Gynecology Hospital (BOGH; 2009–2024), Shijiazhuang Obstetrics and Gynecology Hospital (SOGH; 2018–2020), Xiangya Hospital (XYH; 2022–2024), the Third Affiliated Hospital of Zhengzhou University (TAHZU; 2023–2024), and West China Second Hospital, Sichuan University (WCSHSU; 2023–2025). The ultrasound devices utilized at these centers include: BOGH: GE Voluson E8/E10, Samsung WS80A/HS70A/W10, and Philips EPIQ7; SOGH: GE Voluson E8/E10, Samsung WS80A/RS80A/HS70A, and Philips EPIQ7; XYH: GE Voluson E10, SonoScape P60, and Philips EPIQ7; TAHZU: GE Voluson E8 and Samsung W10; WCSHSU: GE Voluson E8.

### 5.2 Model Configurations and Training

Image-based AI solutions for fetal trisomy 21 screening currently utilize convolutional neural networks (CNNs). We first evaluate the ResNet architecture [20] to establish a baseline for comparative analysis on our datasets. We systematically assess the performance of ResNet under four experimental configurations: normal versus registered ROI cropping, and random initialization versus ImageNet pre-trained weight initialization [21]. This evaluation enables a comprehensive assessment of the impact of data pre-processing and transfer learning.

Building on these findings, we conduct comprehensive pre-training experiments on the registered ultrasound images using the Vision Transformer (ViT) architecture [14].

To rigorously assess the impact of our approach, we evaluate the models across four weight initialization strategies: (1) From scratch (random weight initialization); (2) ImageNet pre-trained (conventional supervised transfer learning); (3) Standard MAE (self-supervised pre-training with 25% random patch masking); and (4) RadarMAE (our sector-masked self-supervised pre-training).

To ensure a fair and equitable comparison across different architectures, we employ the default training configurations and hyperparameters specific to each baseline architecture. For the downstream classification task, we utilize the standard cross-entropy loss function. To mitigate overfitting on the limited fetal ultrasound dataset, we implement standard data augmentation techniques, including random resized cropping and horizontal flipping. Final model selection was determined by the highest Area Under the Curve (AUC) achieved on the internal validation set.

### 5.3 Cross-institutional Generalizability

Cross-institutional deployment of fetal ultrasound AI is challenged by institutionspecific imaging heterogeneity, severe data imbalance, and limited target-domain labels. Differences in ultrasound devices, acquisition protocols, and sonographer expertise cause domain shifts across hospitals, while primary institutions often have too few confirmed abnormal cases to train supervised models independently. In addition, delayed diagnosis, referral, and incomplete follow-up can make reliable imaging labels unavailable in target institutions.

We implemented the Core-and-Branch framework using domain-adversarial neural networks [22]. The core model is trained with labeled data from a central medical center, whereas branch institutions provide only a small number of representative unlabeled images, typically 50–100 cases, for local domain calibration. This design allows diagnostic knowledge learned from data-rich centers to be adapted to target institutions without requiring target-domain diagnostic labels.

As illustrated in Figure 5 (b), the domain-adversarial architecture jointly processes labeled source-domain images and unlabeled target-domain images and optimizes two complementary objectives. The label classifier is trained with source-domain diagnostic labels to preserve trisomy 21 screening performance, whereas the domain classifier is trained through a gradient reversal layer to reduce the distinguishability between source and target features. This adversarial objective encourages the feature extractor to learn domain-invariant fetal facial representations, thereby reducing institution-specific variation without using target-domain diagnostic labels.

### 5.4 Grad-CAM scheme to Quantify Region-Specific Contributions

We adopted Gradient-weighted Class Activation Mapping (Grad-CAM) [23] to quantify region-specific contributions to model predictions by generating saliency heatmaps.

Conventional Grad-CAM processes cases on an individual, image-by-image basis, which prevents the creation of risk maps at the population level and fails to offer macroscopic insights into the condition. As illustrated in Figure 6 (a), our analytical pipeline consists of two steps. In the peak detection step, the global maximum response point is identified as the neural network’s focal locus for each input image. In the spatial aggregation step, all maximum response coordinates across registered images are compiled to generate population-level AI attention maps, thereby shifting the focus from isolated image-level diagnoses to broader, cohort-level insights.

## Data Availability

The clinical imaging data used in this study are not publicly available because they contain sensitive patient-related information. Deidentified data may be made available from the corresponding authors upon reasonable request, subject to approval by the participating institutions and execution of a data-use agreement.

## Acknowledgements

This study was supported by the National Key Research and Development Program of China (Grant Nos. 2022YFC2703300, 2022YFC2703301, and 2023YFC2705602) and the National Natural Science Foundation of China (Grant No. 32370669). The funders had no role in the study design, data collection, data analysis, interpretation of the results, or preparation of the manuscript.

## Data availability

The fetal ultrasound images analysed in this study are not publicly available because they contain sensitive clinical information and are subject to institutional ethics and patient-privacy restrictions. De-identified data may be made available to qualified researchers upon reasonable request to the corresponding authors, subject to approval by the participating institutions and execution of a data-use agreement.

## Code availability

The source code supporting this study is available in the RadarMAE GitHub repository at https://github.com/wujiang5/RadarMAE. During peer review, access to the private repository will be provided to editors and reviewers upon request.

---

**Fig S1.**
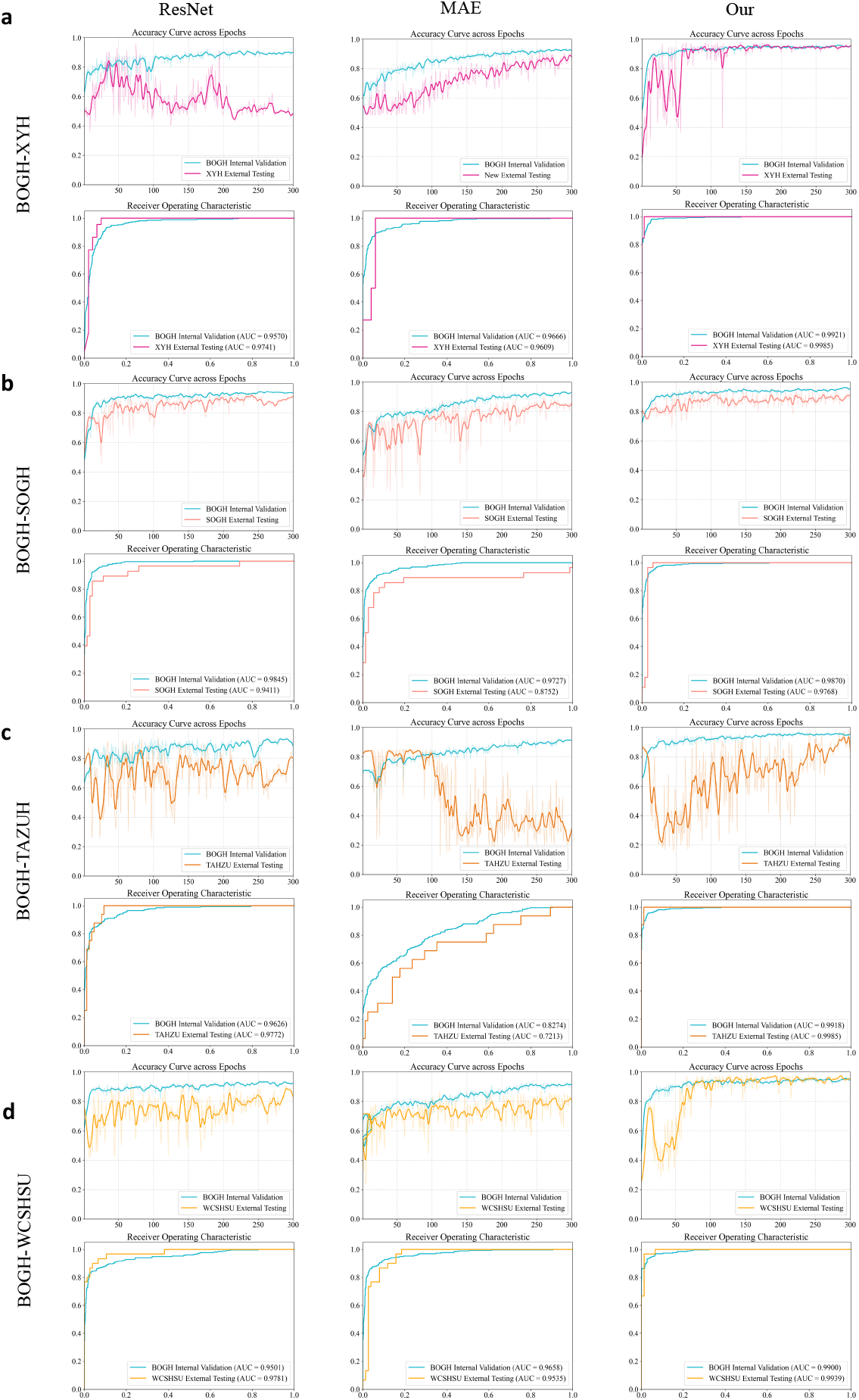
Adversarial training performance between BOGH-XYH, BOGH-SOGH, BOGH-TAHZU, and BOGH-WCSHSU cohort pairs. Accuracy trend and ROC curve using ResNet, MAE, and our RadarMAE as feature extractors in domain-adversarial training. Higher accuracy values and a larger area under the curve indicate a superior performance.

